# Accuracy and confidence calibration of large language models on neurology board-style questions

**DOI:** 10.64898/2026.10.17.26365807

**Authors:** Neil Graham, Stephen Auger, Bahaa Attaallah, Amit Batla, Gregory Scott

**Affiliations:** Applied Health Intelligence Lab, Department of Brain Sciences, Imperial College London, UK; UK Dementia Research Institute Centre for Care Research and Technology, Imperial College London, UK; Department of Targeted Intervention, UCL, UK; Department of Clinical and Movement Neuroscience, UCL Queen Square Institute of Neurology, University College London, UK

**Author notes:** Corresponding Author: Dr Neil Graham MRCP PhD, Applied Health Intelligence Lab, Department of Brain Sciences, Imperial College London, London, United Kingdom.

## Abstract

Large language models (LLMs) are increasingly evaluated for clinical tasks, but safe use requires reliable uncertainty estimates as well as high accuracy. We tested eight general-purpose LLMs on 100 validated neurology board-style multiple-choice questions and compared three confidence signals: self-reported confidence, critique-implied confidence from an independent model instance, and consistency-based confidence from repeated sampling. Accuracy varied from 45% (Llama 3.1 8B) to 92% (Gemini 3 Flash/Pro), but confidence was generally overestimated, particularly in lower-accuracy models. Consistency-based confidence provided the best discrimination between correct and incorrect answers (pooled AUROC 0.74), outperforming self-report, although calibration differences between methods were not significant. Calibration should therefore be reported alongside accuracy when clinical LLMs are evaluated.

**Short Description:** We evaluated the accuracy and confidence of eight large language models on 100 neurology board-style questions. We found substantial overconfidence, and differences between confidence methods in their ability to distinguish correct from incorrect answers.

## INTRODUCTION

Large language models have demonstrated remarkable performance on medical licensing examinations ^1,2^, raising the prospect of their deployment in higher-risk contexts such as clinical decision support or patient consultations. In neurology, where there is a particular shortage of trained specialists^3^ and where much of the diagnostic information is accessible via the history, the potential value of such tools is considerable.

Recognising and working within the limits of your competence is a key duty of a doctor^4^. Escalating issues which fall beyond your knowledge or skill to those with wider or deeper training protects patient safety and means that risk can efficiently be managed within a health system. Hence, both accuracy and calibration (knowing what you know) are critical for clinical safety. A well-calibrated model could undertake useful functions such as triage, answering confidently when likely correct and escalating to a human expert clinician when uncertain. A poorly calibrated model lacks such a safety net and would pose considerable risks.^5^

LLM performance in neurology suggests that recent models may outperform neurologists in vignette-based tasks assessing differential diagnosis.^6^ However, many evaluations of language models in medicine have focused predominantly on accuracy,^2,7^ with limited attention to calibration. In limited work where this has been assessed, models have been found to lack good self-knowledge^8^, although these studies frequently relied on a single method to interrogate model confidence or meta-knowledge.^9–11^ Several approaches have been proposed to assess model confidence, including self-report^12^, self-criticism^13^ and sampling methods that make use of multiple responses.^14^

Here we report a systematic evaluation of eight LLMs on 100 neurology board-style questions, which are benchmarked to the standard of a UK consultant neurologist. These are general purpose models, rather than medically fine-tuned LLMs, reflecting strong priors for their performance in this context,^15^ and a more rapid upgrade cycle. We assessed not only accuracy but also the quality of three distinct measures of confidence: direct self-report, structured critique, and answer consistency across repeated questioning. Our aim was to assess how well model confidence reflected answer correctness and whether this differed by confidence-estimation method.

## METHODS

### Question Bank

From the eBrain interactive learning platform question bank, a set of 100 board-style multiple-choice questions (MCQs) was selected. These questions are mapped to the UK neurology training curriculum and cover a broad range of neurological conditions, including neuromuscular disorders, cerebrovascular disease, neuro-oncology, demyelinating disease, movement disorders, epilepsy, and neuro-ophthalmology. Each question contains a clinical vignette with five answer options and one single best answer. Ninety-three questions were text-only and seven included radiological images alongside the text; these images were not provided to models, as only a proportion of the models evaluated were multi-modal. Ground-truth answers had previously been verified by expert clinicians at the time this validated attending/consultant-level question set was devised. The test is designed to be equivalent in difficulty to the Royal College of Physicians Summary Certificate Examination in neurology, where the typical human passing mark is around 60%. These questions are available only to registered users of the platform and are therefore unlikely to appear in model pre-training data.

### Models

Eight models were selected spanning a range of architectures and sizes. Open-weight models included Meta’s Llama 3.1 at three parameter scales (8 billion, 70 billion, and 405 billion), Moonshot AI’s Kimi K2.5, and DeepSeek’s v3.2. Proprietary models included OpenAI’s GPT-4o, Google’s Gemini 3 Flash and Gemini 3 Pro models. All were accessed via the OpenRouter API to ensure a standardised inference pipeline (via openrouter.ai).

### Experimental Design

Three experiments were conducted for each question, per model. In the direct confidence experiment, each question was presented once at temperature 0.0 (aiming to be more deterministic) and the model was asked to select the correct answer and provide a self-reported confidence score (0-100%). In the second experiment, the critique test, a fresh instance of the same model received the original question together with the first instance’s answer (but not its confidence) and was asked to critically evaluate the answer and estimate the probability that it was incorrect. The critique-implied confidence was computed as 100 minus this estimated error probability. Last, in the consistency experiment, each question was presented 10 times to each model at temperature 0.7. We recorded the modal (most frequent) answer and its selection frequency; for calibration and discrimination analyses of the deterministic answer, consistency-based confidence was defined as the proportion of repeated runs selecting that deterministic answer. Where a tie occurred between two answers in the consistency experiment (10 of 800 model-question pairs), a single additional run of the model was used as a tiebreaker. The same prompt as in experiment 1 was used (See Figure 1). Research ethics committee approval was not required for this analysis of LLMs on MCQ data.

**Figure 1.**
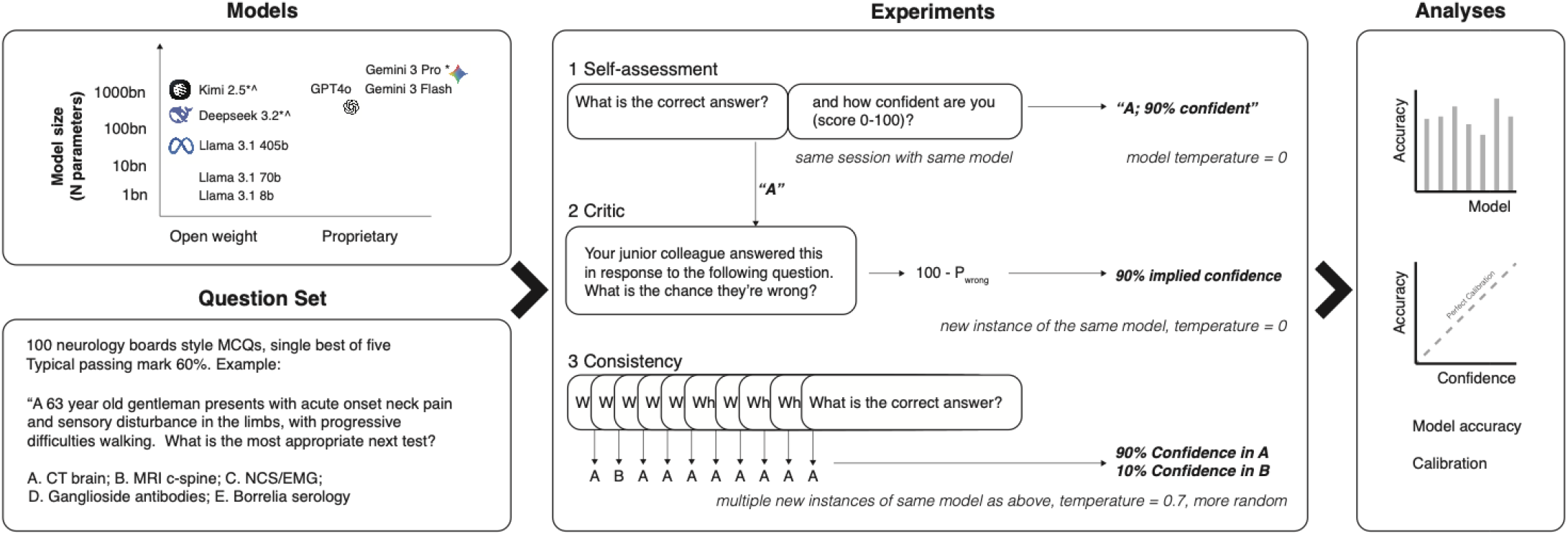
Study Design. 100 board-style neurology questions were presented to eight LLMs. Three confidence elicitationexperiments were conducted. (1) Self-assessment: each model answered the question and provided a confidence rating (0–100%), with temperature set to 0. (2) Critique: a separate instance of the same model (temperature = 0) was told that a juniorcolleague had selected a given answer and asked to estimate the probability of error; implied confidence was computed as100 minus this error estimate. (3) Consistency: ten independent runs of the same model at elevated temperature were used togenerate an answer distribution; confidence in each option was taken as its selection frequency across runs (e.g. 90% foroption A if chosen in 9 of 10 runs). Outputs from all three experiments were then evaluated.

For the deterministic analysis, the following prompt was used: “You are an expert neurologist. Select the single best answer. Output strict JSON: “correct_option”: “<Option Letter>“, “confidence”: <0-100>. Provide your confidence as a probability between 0% and 100%. Note that for a 5-option question, a random guess implies ~20% confidence. Only use 100% if absolutely certain.” For the critique analysis, the following was used: “You are a critical neurology board examiner. Review the candidate’s answer. Estimate the probability (0-100%) that the answer is INCORRECT. Output strict JSON: “probability_incorrect”: <0-100>. Question: <question text> Candidate Answer: step1<correct_option>. Critically evaluate this. What is the probability it is wrong?”

### Statistical Analysis

Accuracy was defined as exact match between the model’s selected option and the ground truth, assessed on the temperature-0 deterministic response. To account for uncertainty in model performance given the sample size of 100 questions, 95% confidence intervals (CIs) were calculated for each model’s accuracy using the Clopper-Pearson method.

Calibration was assessed using expected calibration error (ECE). Predictions were assigned to ten confidence bins: [0,10), [10,20), …, [90,100] with the final bin including confidence values of 100%. Within each bin, the absolute difference between mean predicted confidence and observed accuracy was calculated; ECE was then computed as the weighted mean of these absolute differences, with weights proportional to the number of observations in each bin. Confidence intervals for ECE were estimated using non-parametric percentile bootstrap resampling with 2000 resamples, resampling model-question observations with replacement and recalculating ECE using the same binning scheme.

Discrimination, defined as the ability of a confidence score to distinguish correct from incorrect answers, was assessed using the area under the receiver operating characteristic curve (AUROC). AUROCs and their 95% confidence intervals were estimated using DeLong’s method, implemented in the pROC package. Comparisons between AUROCs for different confidence-estimation methods were also performed using DeLong’s test. Analyses were performed using R (R statistical corp).

## RESULTS

### Model accuracy in multiple choice question answering

Model accuracy in deterministic testing varied from 45% (95% CI 35-55) using the smallest model, Llama 3.1 8B, to 92% (CI 85-96) with Gemini 3 Flash or Gemini 3 Pro (Figure 2, Supplementary Table 1). Within the Llama family, models with greater parameter counts showed greater accuracy, of 45%, 60%, and 71% for 8B, 70B, and 405B variants respectively. The most accurate open-weight model was Kimi K2.5 at 82% (73-89), scoring greater than the human passing mark of 60%.

**Figure 2.**
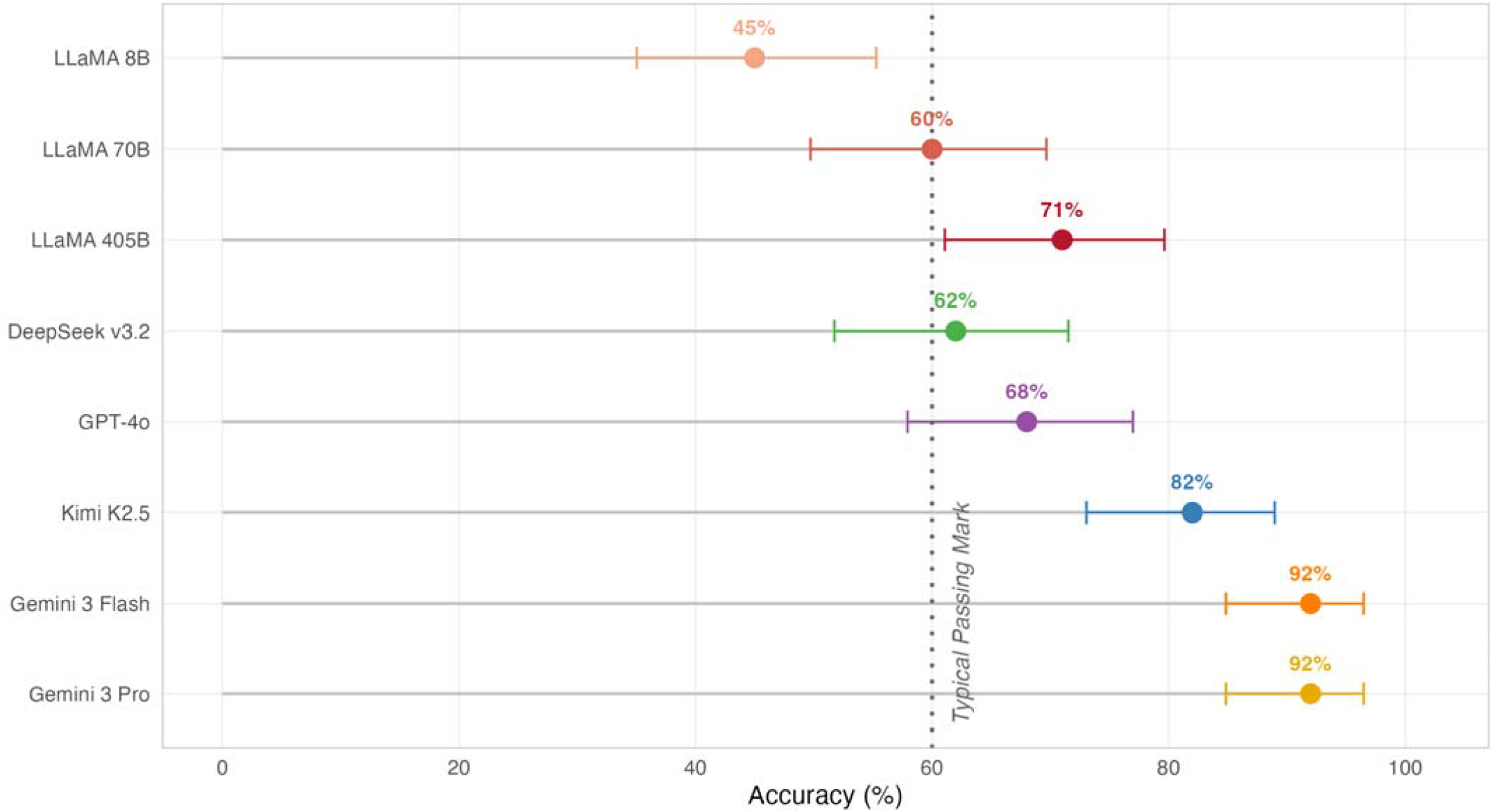
Model Accuracy on Neurology Multiple Choice Questions. Accuracy is represented by the colored points. Horizontal bars indicate 95% exact binomial confidence intervals. The vertical dotted line represents a typical test passing threshold of 60%.

Overall, there was no significant accuracy difference between the single-run (deterministic, temperature 0) and consistency (ten runs, modal answer) approaches, whose answers were the same in n=751 of 800 cases (93.9%). In the disagreement cases, the consistency (modal) answer was more frequently correct. Of note, some text questions referred to images which we did not provide to the models: as expected, LLM accuracy was typically worse on these questions (mean accuracy of 55.4%, 41.5% - 68.7%) than text-only questions (72.7%, 69.4% - 75.9%).

### Calibration and discrimination

Confidence was poorly calibrated across all models (Table 1, Figure 3) with LLMs overconfident on average. Expected calibration error (ECE) was calculated, representing deviation from perfect calibration (i.e., lower values are better), which is not affected by averaging-out effects where there is mixed over and under-confidence. The critique-implied method had the lowest ECE with a mean of 17.8 (SD 11.6), followed by self-reported (18.5, SD 13.0) and last by consistency (mean 20.0, SD 9.2), though these differences were not statistically significant (Friedman test p>0.05). Kimi K2.5 and Gemini 3 Flash had the lowest ECEs (of 3.9 and 4.7 respectively) with Llama 8B and 70B poorest performers (ECEs of 30.6, 35.3).

**Table 1.** Language model accuracy and discrimination. For expected calibration error, lower scores reflect better calibration); AUROC = area under the receiver operating characteristic curve (higher indicates better discrimination between correct and incorrect answers). Accuracy figures reflect deterministic (temperature 0testing. Each cell reports the point estimate followed by a 95% confidence interval in parentheses: Clopper-Pearson exact binomial CI for accuracy, percentile bootstrap (B = 2000) CI for ECE, and DeLong’s CI for AUROC.

| Model | Accuracy,<br>(95% CI) | Expected Calibration Error<br>(95% CI) |  |  | Discrimination (AUROC)<br>(95% CI) |  |  |
| --- | --- | --- | --- | --- | --- | --- | --- |
|  |  | Self | Critique | Consistency | Self | Critique | Consistency |
| LLaMA 8B | 45.0 (35.0-55.3) | 42.3 (33.1-51.9) | 30.6 (20.8-41.0) | 26.1 (19.8-36.1) | 0.470 (0.364-0.576) | 0.521 (0.430-0.612) | 0.697 (0.595-0.799) |
| LLaMA 70B | 60.0 (49.7-69.7) | 26.9 (17.7-36.7) | 35.3 (25.8-45.4) | 29.3 (21.9-39.0) | 0.555 (0.452-0.659) | 0.555 (0.472-0.638) | 0.680 (0.585-0.776) |
| LLaMA 405B | 71.0 (61.1-79.6) | 11.8 (3.7-20.9) | 23.5 (14.8-32.5) | 25.0 (17.4-34.2) | 0.614 (0.528-0.701) | 0.571 (0.456-0.686) | 0.640 (0.539-0.741) |
| DeepSeek v3.2 | 62.0 (51.7-71.5) | 26.6 (17.3-35.7) | 18.1 (12.1-27.9) | 21.5 (15.2-29.4) | 0.718 (0.622-0.815) | 0.720 (0.617-0.823) | 0.827 (0.745-0.910) |
| GPT-4o | 68.0 (57.9-77.0) | 20.4 (11.4-29.2) | 16.8 (9.4-25.2) | 29.6 (21.8-39.3) | 0.618 (0.509-0.727) | 0.763 (0.672-0.854) | 0.617 (0.534-0.700) |
| Kimi K2.5 | 82.0 (73.1-89.0) | 10.4 (4.6-18.2) | 3.9 (2.3-10.5) | 13.7 (8.1-20.7) | 0.767 (0.657-0.877) | 0.876 (0.786-0.967) | 0.806 (0.690-0.922) |
| Gemini 3 Flash | 92.0 (84.8-96.5) | 5.5 (1.6-10.6) | 4.7 (1.1-10.2) | 8.3 (3.8-14.1) | 0.826 (0.682-0.970) | 0.774 (0.587-0.960) | 0.535 (0.411-0.660) |
| Gemini 3 Pro | 92.0 (84.8-96.5) | 4.4 (0.3-10.1) | 9.5 (4.4-15.7) | 6.7 (2.7-11.7) | 0.714 (0.623-0.805) | 0.584 (0.425-0.744) | 0.738 (0.553-0.924) |

**Figure 3.**
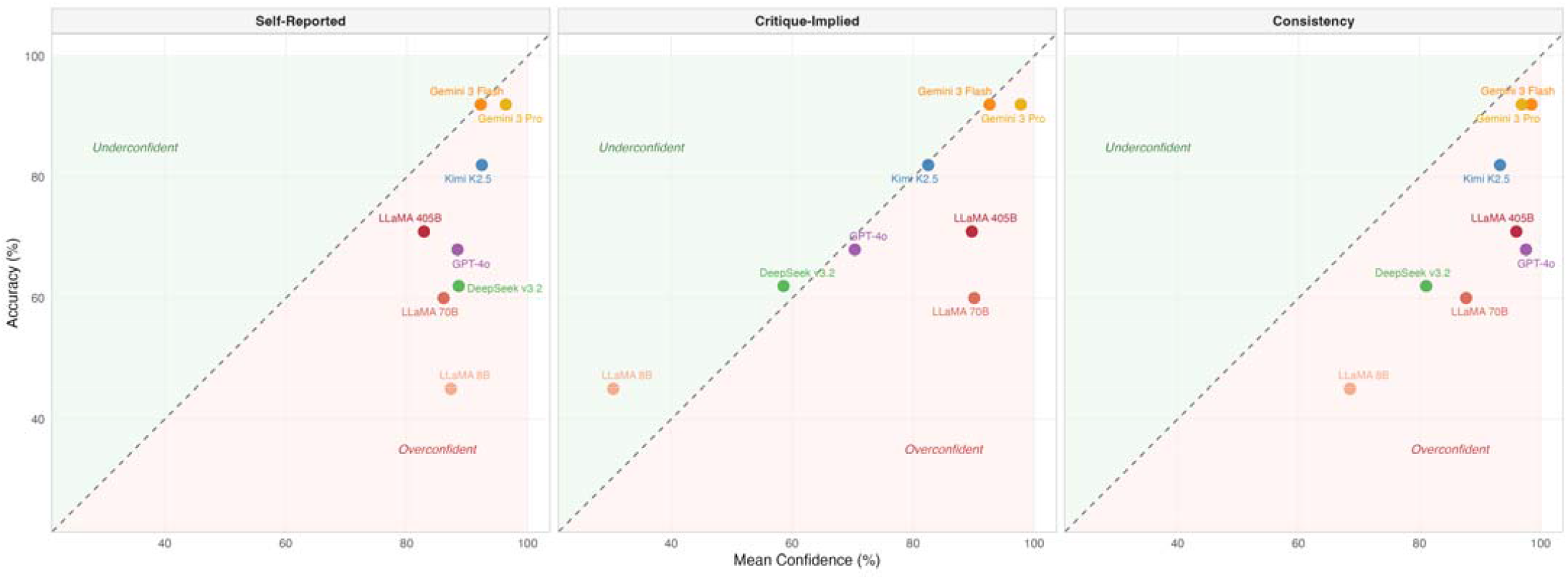
Model Accuracy vs Confidence. Each point represents a language model. The dashed diagonal line represents perfect calibration (confidence equals accuracy) with points below this indicating overconfidence and points above underconfidence. Left panel: self-reported confidence; centre: critique-implied confidence; right: consistency-based confidence. All models fall in the overconfident region for self-reported confidence, with smaller models (Llama 8B) furthest from the diagonal.

Next, the ability of model confidence outputs to discriminate right vs wrong answers was tested with AUROCs calculated. Across all the models, the best-performing approach was consistency with an AUROC of 0.74, numerically greater than the critique-implied method (0.70), and self-report (0.69). Consistency was significantly better than self-report (DeLong’s test p=0.048), but other comparisons did not meet this statistical threshold. The top performing model, looking across all methods, was Kimi K2.5 (AUROC 0.88, using critique-implied confidence) followed by Gemini 3 Flash (0.83, on self-report) with the smallest model tested performing worse than chance (0.47; Llama 8B, self-report).

## DISCUSSION

Here, we show that large frontier models demonstrate high levels of accuracy on challenging specialist board-style neurology questions, often greater than the human passing score (60%). However, this varied widely with the LLM examined (from 45–92%), with clear scaling effects as larger models perform better. Importantly, we found that modern large open weight models performed above the typical passing threshold of 60%, in respect of both Llama 3.1 405B (71%) and Kimi K2.5 (82%), suggesting the clinical potential of locally deployable privacy-preserving models. Our accuracy data extend prior evaluations of LLMs on US neurology board examinations, suggesting that newly developed models can achieve even greater accuracy (>90%) than the strongest performers in prior assessments^16^.

However, we found the ability of models to attach reliable confidence estimates to their assertions was limited, tending towards overconfidence, and being especially misleading for lower-accuracy models, which are precisely those most in need of reliable uncertainty signaling. This mirrors work outside of medical domains suggesting Dunning-Kruger-like behaviour by LLMs^17^. In addition to self-reported confidence, two further approaches were evaluated which might have produced better calibration: opening a new model instance and critically evaluating the previous response (which might reduce ‘anchoring bias’) or a ‘consistency approach’, sampling from the model’s probability distribution by taking the modal answer across ten runs. These alternative approaches shifted the confidence landscape although there was no statistically significant difference in calibration when comparing approaches across all of the models.

Our work suggests that the optimal confidence strategy may be model-dependent, with poor calibration potentially reflecting a fundamental challenge of model development strategies such as reinforcement learning with human feedback, which may select for answers which are more definitive^18^. Notably, the low temperature ‘deterministic’ answer differed from the consistency answer on some occasions, suggesting that the globally most probable response was not necessarily retrieved by selecting the most likely next token. This may relate to a requirement to sequentially select several tokens in turn within this task, and highlights that selecting a temperature of 0 may not fully remove stochastic behaviour, due to rounding errors and batch-processing. This is important for healthcare developers to note in developing LLM-based tools where reproducibility is required.

Cost-effectiveness is a further important criterion for clinical deployment. We observed similar performance between Google’s frontier reasoning model, Gemini 3 Pro, and the smaller/faster Gemini 3 Flash model. Both were 92% accurate and had similar calibration. Because lower-cost models may be attractive for clinical deployment, this finding suggests that model selection should consider calibration and accuracy alongside cost. It is unclear whether similar performance reflects a ceiling of this model architecture on the benchmark or whether the two models are genuinely similar in this use case. Notably, cost per unit of intelligence has fallen substantially over time, with token costs for a GPT-4-class model estimated to have reduced 1000-fold from 2023 to 2025.^20^

There are several important limitations. The clinical vignette format, while standard in these examinations, does not capture the complexity of clinical reasoning in practice, and future work will be needed to assess accuracy and calibration in settings bearing greater similarity to the clinic. The question bank of 100 items, while covering a broad range of neurological topics, is modest in size; we did not provide images to multi-modal models, and we did not evaluate all contemporary model families (e.g., Grok, Claude). Finally, our study evaluates models in isolation rather than in human-AI collaborative settings, where the interaction between model confidence and clinician behaviour may introduce additional complexity.

In conclusion, our data indicate that accuracy alone is an insufficient metric for evaluating clinical LLMs. Confidence-based abstention is a potential application of uncertainty estimates,^19^ but selecting and validating an escalation rule would require separate data and evaluation in clinical settings. We recommend that calibration be reported as a standard evaluation axis, and that future work in this domain explore in more detail how this information, alongside accuracy, affects clinical human-AI teaming.

## FUNDING AND ACKNOWLEDGEMENTS

NG acknowledges funding support for this work from the NIHR via an Advanced Fellowship (NIHR306582), SA acknowledges a Clinical Lectureship from NIHR, and GS acknowledges an Advanced Fellowship from the NIHR. NG, GS, and SA acknowledge the support of the UK Dementia Research Institute Centre for Care Research and Technology, and the Imperial NIHR BRC. AB has been funded by MRC-NIHR, MSA Trust, and UCL Centre for Equality Research in Brain Sciences. We acknowledge the support of Elinor and Adrian Wills from the Joint Neurosciences Council. The funders had no role in study design, data collection, analysis, interpretation, manuscript preparation, or the decision to submit.

## AUTHOR CONTRIBUTIONS

NG conceived the study, designed the analysis, performed the experiments and statistical analysis, and drafted the manuscript. SA and GS contributed to study design, interpretation of results, and manuscript revision. BA and AB contributed clinical expertise, interpretation of results, and manuscript revision. All authors reviewed and approved the final manuscript. Use of AI: GPT5 and 6 (Astra; OpenAI) were used to assist in R coding for the data analyses and to improve the grammar/brevity of the manuscript when drafted.

## DATA AND CODE AVAILABILITY

Data and analysis scripts for the manuscript are available via zenodo and github respectively. Confidence and calibration data: https://doi.org/10.5281/zenodo.20272032 and interactive python R notebook for analyses: https://github.com/Applied-Health-Intelligence-Lab/Neurology_LLM_Confidence_Calibration/tree/main. The eBrain question text, answer options, and answer key are not publicly available because the questions are restricted educational assessment materials and release would compromise assessment integrity; access is controlled by the eBrain platform.

## SUPPLEMENTARY MATERIALS

### SUPPLEMENTARY RESULTS

**Supplementary Table 1.**
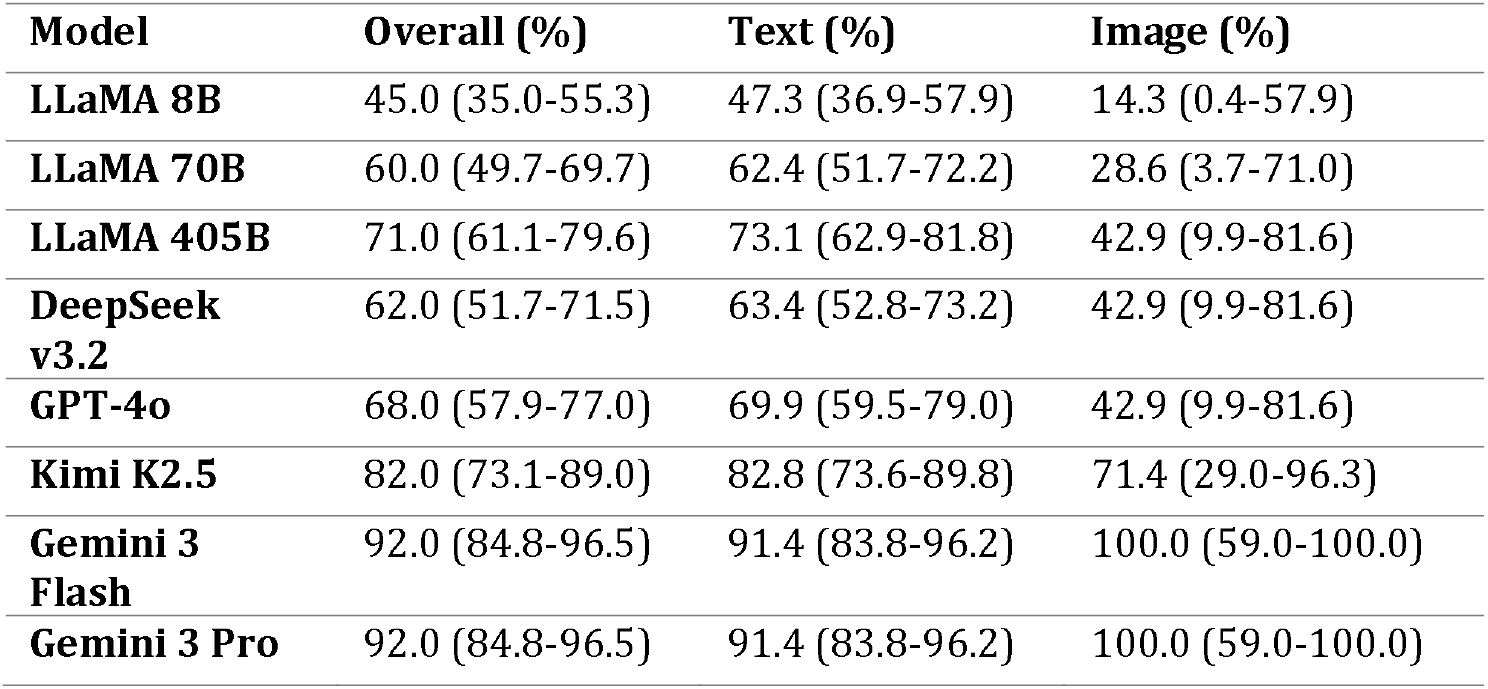
Accuracy of Eight Large Language Models on Image versus Text Questions. Text questions, n=93; Image questions, n=7. Models are ordered by overall accuracy. Deterministic testing approach used.

| Model | Overall (%) | Text (%) | Image (%) |
| --- | --- | --- | --- |
| **LLaMA 8B** | 45.0 (35.0-55.3) | 47.3 (36.9-57.9) | 14.3 (0.4-57.9) |
| **LLaMA 70B** | 60.0 (49.7-69.7) | 62.4 (51.7-72.2) | 28.6 (3.7-71.0) |
| **LLaMA 405B** | 71.0 (61.1-79.6) | 73.1 (62.9-81.8) | 42.9 (9.9-81.6) |
| **DeepSeek v3.2** | 62.0 (51.7-71.5) | 63.4 (52.8-73.2) | 42.9 (9.9-81.6) |
| **GPT-4o** | 68.0 (57.9-77.0) | 69.9 (59.5-79.0) | 42.9 (9.9-81.6) |
| **Kimi K2.5** | 82.0 (73.1-89.0) | 82.8 (73.6-89.8) | 71.4 (29.0-96.3) |
| **Gemini 3 Flash** | 92.0 (84.8-96.5) | 91.4 (83.8-96.2) | 100.0 (59.0-100.0) |
| **Gemini 3 Pro** | 92.0 (84.8-96.5) | 91.4 (83.8-96.2) | 100.0 (59.0-100.0) |

## REFERENCES

1 Singhal, K. et al. Large language models encode clinical knowledge. Nature 620, 172–180 (2023). 10.1038/s41586-023-06291-2

2 Nori, H., King, N., McKinney, S. M., Carignan, D. & Horvitz, E. Capabilities of GPT-4 on medical challenge problems. arXiv 10.48550/arXiv.2303.13375 (2023).

3 Association of British Neurologists. The State of the UK Neurology Workforce: Needs for a Population of 100,000 (Association of British Neurologists, 2026). https://www.theabn.org/page/workforce

4 General Medical Council. Good medical practice (General Medical Council, 2024). https://www.gmc-uk.org/professional-standards/professional-standards-for-doctors/good-medical-practice

5 Guo, C., Pleiss, G., Sun, Y. & Weinberger, K. Q. On calibration of modern neural networks. In Proceedings of the 34th International Conference on Machine Learning, vol. 70, 1321–1330 (PMLR, 2017).

6 Barrit, S. et al. Specialized large language model outperforms neurologists at complex diagnosis in blinded case-based evaluation. Brain Sci. 15, 347 (2025). 10.3390/brainsci15040347

7 Kung, T. H. et al. Performance of ChatGPT on USMLE: potential for AI-assisted medical education using large language models. PLOS Digit. Health 2, e0000198 (2023). 10.1371/journal.pdig.0000198

8 Bentegeac, R., Le Guellec, B., Kuchcinski, G., Amouyel, P. & Hamroun, A. Token probabilities to mitigate large language models overconfidence in answering medical questions: quantitative study. J. Med. Internet Res. 27, e64348 (2025). 10.2196/64348

9 Omar, M., Agbareia, R., Glicksberg, B. S., Nadkarni, G. N. & Klang, E. Benchmarking the confidence of large language models in answering clinical questions: cross-sectional evaluation study. JMIR Med. Inform. 13, e66917 (2025). 10.2196/66917

10 Almomani, M. et al. Evaluation of multiple generative large language models on neurology board-style questions. Front. Digit. Health 7, 1737882 (2026). 10.3389/fdgth.2025.1737882

11 Savage, T. et al. Large language model uncertainty proxies: discrimination and calibration for medical diagnosis and treatment. J. Am. Med. Inform. Assoc. 32, 139–149 (2025). 10.1093/jamia/ocae254

12 Tian, K. et al. Just ask for calibration: strategies for eliciting calibrated confidence scores from language models fine-tuned with human feedback. In Proceedings of the 2023 Conference on Empirical Methods in Natural Language Processing, 5433–5442 (Association for Computational Linguistics, 2023). 10.18653/v1/2023.emnlp-main.330

13 Kadavath, S. et al. Language models (mostly) know what they know. arXiv 10.48550/arXiv.2207.05221 (2022).

14 Xiong, M. et al. Can LLMs express their uncertainty? An empirical evaluation of confidence elicitation in LLMs. In International Conference on Learning Representations (ICLR, 2024). 10.48550/arXiv.2306.13063

15 Nori, H. et al. Can generalist foundation models outcompete special-purpose tuning? Case study in medicine. arXiv 10.48550/arXiv.2311.16452 (2023).

16 Schubert, M. C., Wick, W. & Venkataramani, V. Performance of large language models on a neurology board-style examination. JAMA Netw. Open 6, e2346721 (2023). 10.1001/jamanetworkopen.2023.46721

17 Qazi, I. A. et al. Large language models show Dunning-Kruger-like effects in multilingual fact-checking. Sci. Rep. 16, 7594 (2026). 10.1038/s41598-026-39046-w

18 Leng, J., Huang, C., Zhu, B. & Huang, J. Taming overconfidence in LLMs: reward calibration in RLHF. arXiv 10.48550/arXiv.2410.09724 (2024).

19 Geifman, Y. & El-Yaniv, R. Selective classification for deep neural networks. Adv. Neural Inf. Process. Syst. 30 (2017).

20 Demirer, M., Fradkin, A., Tadelis, N. & Peng, S. The emerging market for intelligence: pricing, supply, and demand for LLMs. National Bureau of Economic Research Working Paper No. 34608 (2025). 10.3386/w34608

